# SARS-CoV-2 Evolution in an HIV-Endemic Setting: A Genomic Epidemiology Study from Botswana

**DOI:** 10.1101/2025.11.21.25340589

**Authors:** Xuhua Geng, Ivan Barilar, Monamodi Kesamang, Kefentse Arnold Tumedi, Maitshwarelo Matsheka, Chawangwa Modongo, Volodymyr Minin, Stefan Niemann, Sanghyuk S. Shin

## Abstract

**Background:** HIV infection can affect SARS-CoV-2 infection dynamics through prolonged viral replication and altered immune responses, but population-level genomic evidence from HIV-endemic settings is limited. Botswana, a country with high HIV prevalence and an established genomic surveillance infrastructure, provides an important setting to examine how host immune variation influences viral evolution during the Omicron period.

**Methods:** We conducted a population-based genomic epidemiology study in the Greater Gaborone District between June 2022 and November 2024. Among 404 SARS-CoV-2 positive samples, 387 yielded high-quality genomes (Ct < 35). Phylogenetic trees were reconstructed using IQ-TREE, time-calibrated with TreeTime, and visualized in Nextstrain to illustrate lineage turnover and clustering patterns. Participant metadata, including HIV status and vaccination history, were summarized descriptively and incorporated into phylogenetic visualizations to annotate sequences by HIV status.

**Results:** Six successive Omicron sublineage waves (BA.4/BA.5, BQ/FN, XBB, BA.2.86, KP, and XEC.9) were detected, consistent with repeated viral introductions rather than sustained local diversification. Sequences from HIV-positive and HIV-negative individuals were broadly interspersed throughout the phylogeny, indicating no population-level separation by HIV status. However, small dyads of HIV-positive hosts appeared within BA, JN, and KP sublineages.

**Interpretation:** In a highly treated and vaccinated population, HIV infection did not fundamentally alter SARS-CoV-2 evolutionary trajectories at the population level. However, subtle clustering among HIV-positive hosts suggests possible host-specific effects, underscoring the importance of continued genomic surveillance integrating host immunologic and clinical data to detect early signals of viral adaptation.

## Introduction

SARS-CoV-2 has continued to evolve into genetically and antigenically distinct lineages since its emergence, driving successive epidemic waves worldwide ^1^. In particular, the Omicron period has been marked by accelerated viral diversification, with frequent emergence and replacement of sublineages such as BA.4, BA.5, BQ, XBB, and BA.2.86 that have successively shaped global epidemic trajectories^2^. Characterizing how these global evolutionary dynamics are reflected at the regional level is essential for understanding local epidemic trajectories and informing genomic surveillance efforts. Although several studies have documented the global spread and diversification of Omicron sublineages^3,4,5^, and large-scale sequencing consortia and global genomic platforms, such as the COVID-19 Genomics UK (COG-UK) Consortium, the Global Initiative on Sharing All Influenza Data (GISAID), and Nextstrain, have facilitated worldwide phylogenetic analyses and mapping of SARS-CoV-2 lineages across continents^6^, region-specific genomic analyses remain critical to elucidate local transmission dynamics, evolutionary pressures, and contextual factors such as host comorbidities.

In Africa, where Human Immunodeficiency Virus (HIV) prevalence remains high^7^, structural constraints in health-care delivery (e.g., uneven antiretroviral therapy (ART) coverage and viral-suppression, service disruptions) and limited pathogen-genomic surveillance capacity^8,9^ can sustain prolonged infections and create conditions conducive to within-host viral diversification, potentially facilitating the emergence of novel variants^10,11^. However, despite the dual burden of HIV and COVID-19 in Africa^12,13^, population-based genomic data from the region remain limited^14,15^.

Within this broader context, Botswana represents a particularly important setting for understanding the interaction between SARS-CoV-2 evolution and host immune status. Earlier epidemic waves in the country were dominated by the Beta and Omicron variants^13^, consistent with regional patterns observed across southern Africa^3^, but the evolutionary dynamics during 2022-2024, when successive Omicron sublineages replaced one another globally, remain poorly characterized. Moreover, it is unclear whether HIV infection has influenced local transmission patterns or contributed to distinct phylogenetic clustering. Addressing this gap is essential for contextualizing variant emergence in regions where HIV prevalence remains among the highest worldwide.

This study aimed to characterize the temporal and phylogenetic patterns of SARS-CoV-2 Omicron lineage replacement in Botswana between mid-2022 and late 2024, and to assess whether HIV infection influenced these transmission dynamics. To address this, we conducted a population-based genomic epidemiology study in the Greater Gaborone District of Botswana. Between June 2022 and November 2024, sequencing 404 SARS-CoV-2 genomes (387 high-quality) and integrating demographic, clinical, and HIV status information. We reconstructed time-scaled phylogenies and characterized lineage turnover to elucidate local evolutionary dynamics and evaluate whether HIV infection influenced viral transmission patterns.

## Method

### Study population and sample collection

Participants were recruited through enhanced health facility-based surveillance, including screening of clinic attendees for SARS-CoV-2 using a rapid diagnostic test, as well as passive diagnosis of symptomatic cases and contact tracing in five neighborhoods of Greater Gaborone. Only individuals with reverse transcription polymerase chain reaction (RT-PCR) – confirmed SARS-CoV-2 infection were eligible. Demographic, clinical, HIV status, and COVID-19 vaccination data were obtained through standardized questionnaires and electronic health records. Participants with unknown HIV status were offered rapid testing, and those with HIV infection were further characterized by CD4 count, viral load, and ART history. The study protocol was approved by the Health Research and Development Committee (HRDC) of Botswana and by the Institutional Review Board of the University of California, Irvine. Written informed consent was obtained from all participants. For children <7 years, parental/guardian consent was obtained; for children aged 7–17 years, both parental/guardian consent and child assent were obtained.

### Sample processing and sequencing

Nasal swabs were processed at the Botswana Ministry of Health and Wellness National Health Laboratory. Viral RNA was extracted using the QIAamp Viral RNA kit (QIAGEN, Hilden, Germany), and samples with Ct < 35 were processed for sequencing. Whole-genome sequencing was performed using the ARTIC tiled amplicon protocol. Viral genomes were sequenced either on the MinION platform (Oxford Nanopore Technologies, UK) at the Botswana Institute of Technology Research and Innovation or on Illumina platforms (iSeq, Illumina Inc., San Diego, CA, USA) at the Research Center Borstel, Germany. Consensus genomes were generated using the ARTIC bioinformatics pipeline, and downstream analyses were performed using a modified version of V-pipe.

### Lineage assignment and phylogenetic analysis

A total of 404 genomes with associated metadata were generated. Sequences with <80% genome coverage of the Wuhan-Hu-1 reference were excluded, leaving 387 genomes for downstream analyses. Clade assignments and quality assessments were performed using Nextclade (v2.14.0). Phylogenetic reconstruction was carried out with the Nextstrain build workflow (Snakemake v9.8.1): maximum-likelihood trees were inferred with IQ-TREE (v3.0.1) under the General Time Reversible (GTR) substitution model, and time-resolved phylogenies were estimated with TreeTime (v0.10.1).

### Data visualization and downstream analyses

Phylogenies were exported as Auspice JSON for interactive visualization in Nextstrain (Auspice v2.63.0). Additional analyses were performed in R using the ape (v5.8.1), ggtree (v3.14.0), and related packages. Newick trees and Auspice JSON files were parsed to extract node-specific sampling times and reconstruct time-scaled subtrees. Branch lengths were recalibrated to calendar time using sampling dates. Subtrees for major lineages (e.g., BA, BE, BQ, XBB) were generated and exported in Newick format for downstream analyses.

## Results

### Demographic characteristics of participants

A total of 387 participants were included in the analysis, among whom 278 were HIV-negative, 76 were HIV-positive, and 33 had unknown HIV status (Table 1). The mean age of participants was 36.79 ± 13.69 years among HIV-negative individuals, 43.38 ± 11.27 years among those with HIV infection, and 33.06 ± 14.92 years among those with unknown status (*p* < 0.0001). Females accounted for 68.0%, 78.9%, and 51.5% of participants in the respective groups (*p* = 0.069). COVID-19 vaccination coverage was high across all groups ≥87.9%), and no significant difference was observed by HIV status (*p* = 0.572). Most participants reported no recent travel (≥83.5%), with a small proportion indicating domestic or international travel (*p* = 0.275). Overall, the demographic composition was broadly comparable across HIV strata, except for age, which was significantly higher among HIV - positive participants.

**Table 1.**
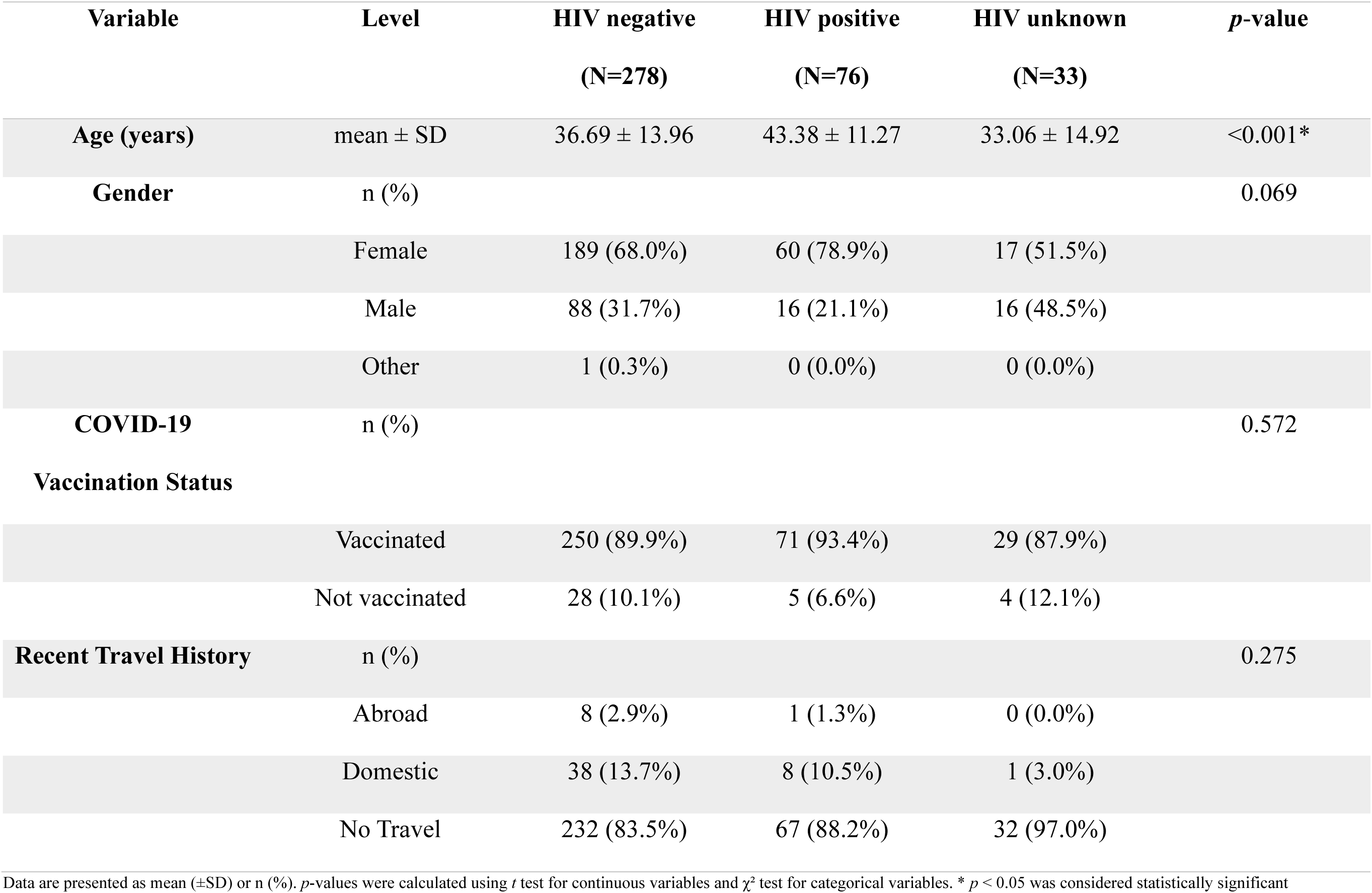
Demographic characteristics of participants stratified by HIV status.

### Epidemiological dynamics

We analyzed 387 high-quality SARS-CoV-2 genomes collected from the Greater Gaborone District, Botswana, between June 2022 and November 2024. The temporal distribution of parent lineages is summarized in Figure 1. We observed several distinct replacement patterns of specific sublineages over the study period. BA sublineages (predominantly BA.4 and BA.5) circulated widely from June through October 2022. These were subsequently replaced by BQ (primarily BQ.1.1) alongside co-circulating FN lineages during November 2022 to January 2023. From May to September 2023, XBB lineages became the dominant variant. In October 2023, the BA.2.86 clade and its sublineages (e.g., BA.2.86.2, BA.2.86.3, and BA.2.86.7) displaced XBB and remained predominant until April 2024. Between May and July 2024, KP lineages expanded rapidly, with KP.1.1 accounting for the majority of detections and additional identification of KP.2, KP.2.2, and KP.2.3. XEC was first detected in June 2024 and subsequently expanded, with XEC.9 becoming the dominant lineage by October 2024, followed by a modest resurgence of KP in November 2024. Overall, these findings reveal distinct sequential waves of lineage turnover in Botswana: BA.4/BA.5 (mid-2022), BQ/FN (late 2022 to early 2023), XBB (mid-2023), BA.2.86 (late 2023 to early 2024), KP (mid-2024), and XEC.9 (late 2024).

**Figure 1:**
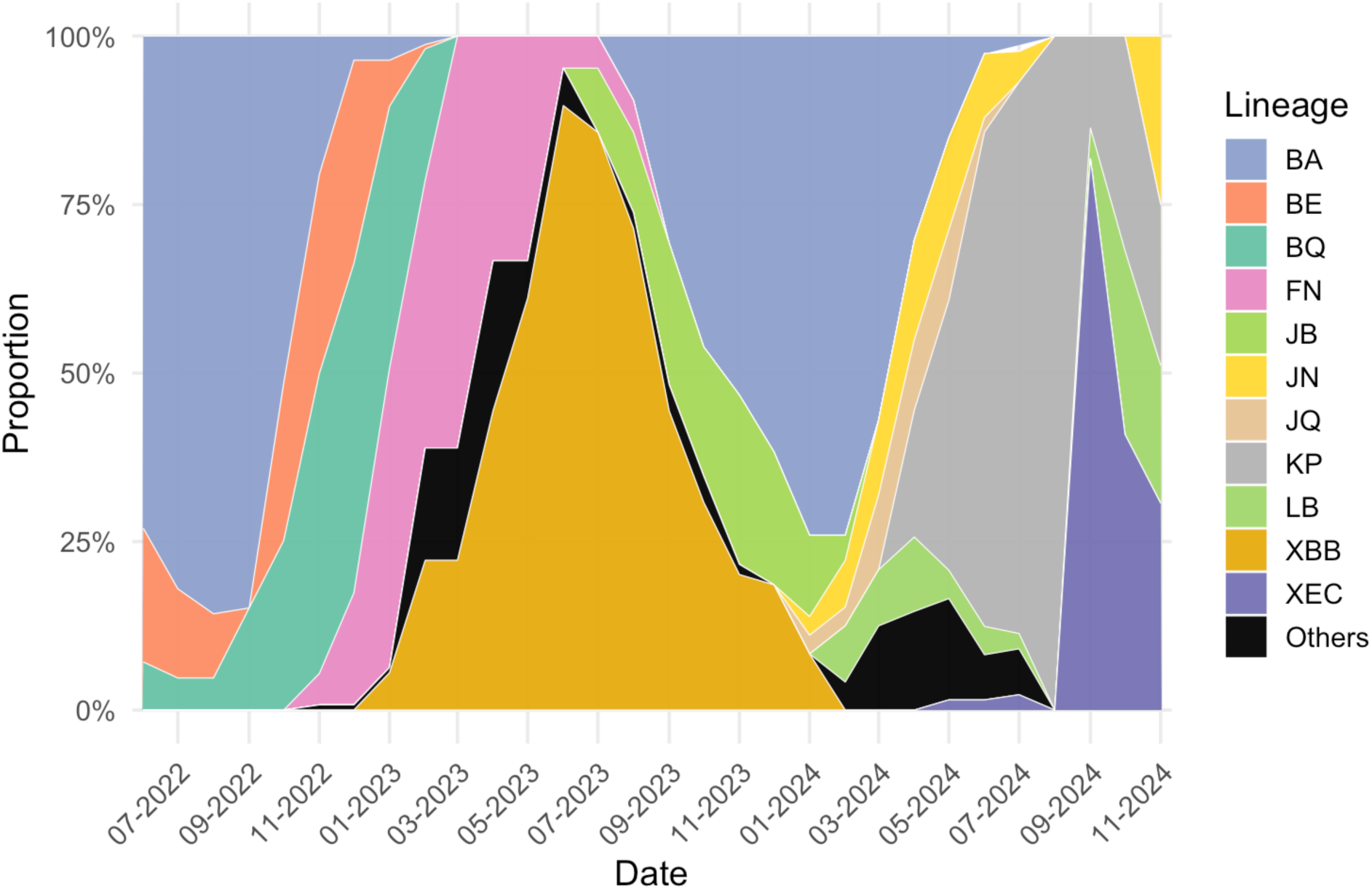
Temporal distribution of SARS-CoV-2 lineages in Botswana, June 2022–November 2024. Stacked area plot showing the monthly proportion of parent lineages among 387 high-quality genomes collected in the Greater Gaborone District. Parent categories (e.g., BA, XBB, KP) are displayed, while specific sublineages (e.g., BA.4, BA.5, BQ.1.1, BA.2.86.2, KP.2.3, XEC.9) were identified from Pango lineage assignments in the sequencing metadata. Three major waves of lineage turnover were observed: BA.4/BA.5 predominated in mid-2022, followed by BQ/FN and XBB in late 2022–2023, and subsequently replaced by BA.2.86, KP, and XEC.9 during 2024.

### Phylogenetic analysis

The phylogenetic analysis in Figure 2 further resolves the evolutionary relationships of the lineages and clarifies the origins of these dynamics within Botswana. Globally, BA.4 (22A) and BA.5 (22B) formed sister lineages with a shared ancestor, and BA.5 diversified further to seed BQ.1 (22E). In Botswana, BA.4 and BA.5 dominated during mid-2022, consistent with global trends. BQ.1 subsequently expanded and surpassed BA.5 in cluster size, although its dominance was transient and it was eventually displaced by XBB and BA.2.86/JN.1 lineages. XBB lineages (23A, 23B, 23D) emerged globally as recombinants of BA.2 sublineages. In Botswana, XBB variants were first detected in early 2023, but sequences were scattered across multiple subclades, including XBB.1.5 and XBB.1.16, rather than forming a monophyletic cluster. This pattern is consistent with repeated introductions from abroad rather than sustained local diversification. The phylogeny also delineates the emergence of BA.2.86 (23I) and its descendant JN.1 (24A). The estimated time to the most recent common ancestor (tMRCA) of BA.2.86 falls between September 2022 and March 2023; however, Botswana sequences were only detected thereafter, suggesting an external origin and subsequent importation. JN.1 was sporadically detected in Botswana as early as March 2023, but broader circulation, including the emergence of sublineages such as 24B, 24C, and 24F, was established in early 2024 and expanded rapidly throughout 2024.

**Figure 2:**
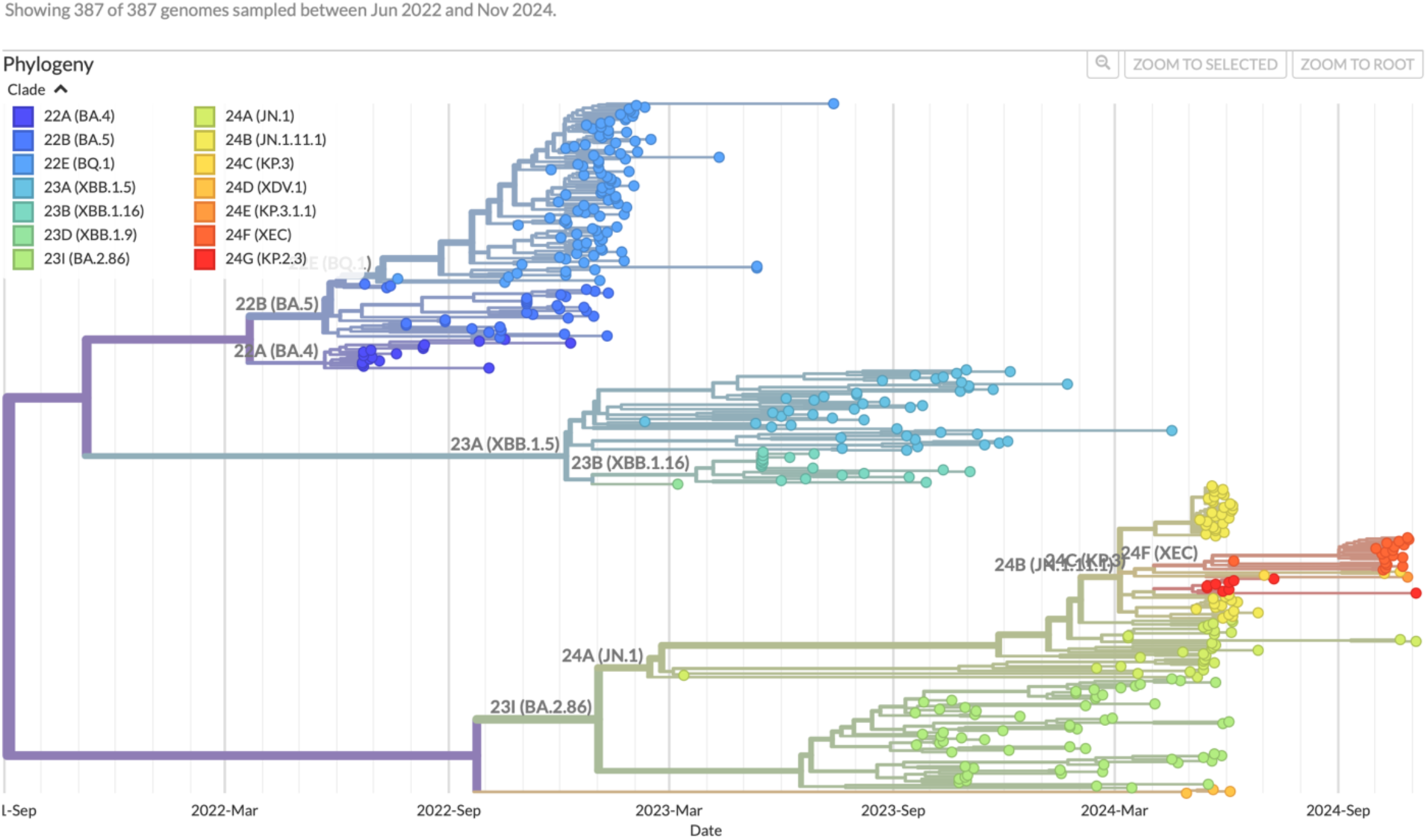
Time-resolved phylogeny of 387 SARS-CoV-2 genomes from Botswana. Maximum-likelihood phylogeny annotated with Nextstrain clade designations, showing sequential replacement of lineages between June 2022 and November 2024. Early genomes clustered within BA.4 (22A) and BA.5 (22B) clades, followed by diversification into BQ.1 (22E) and XBB (23A, 23B, 23D). A third wave began in late 2023 with the emergence of BA.2.86 (23I) and JN.1 (24A), which diversified into descendant clades in 2024, including 24B (JN.1.1.1), 24C (KP.3), and 24E (KP.1.1). The most recent tips were dominated by XEC (24F), particularly XEC.9, along with detections of KP.2.3 (24G), reflecting the ongoing diversification of JN.1-derived lineages.

In addition to the overall phylogeny, we examined lineage-specific time-scaled trees annotated by HIV status to evaluate whether HIV infection structured transmission patterns (Figure 3). Across all major lineages (BA, BE, BQ, FN, JB, JN, JQ, KP, LB, XBB, and XEC), sequences from HIV - positive and HIV - negative individuals were broadly interspersed. Occasional sub-branches, including dyads of HIV - positive hosts, were observed within BA, JN, and KP but remained limited in scale and did not constitute well-defined clusters. Two lineages with small sample sizes, BE (n = 19) and LB (n = 10), comprised only HIV-negative or unknown hosts.

**Figure 3:**
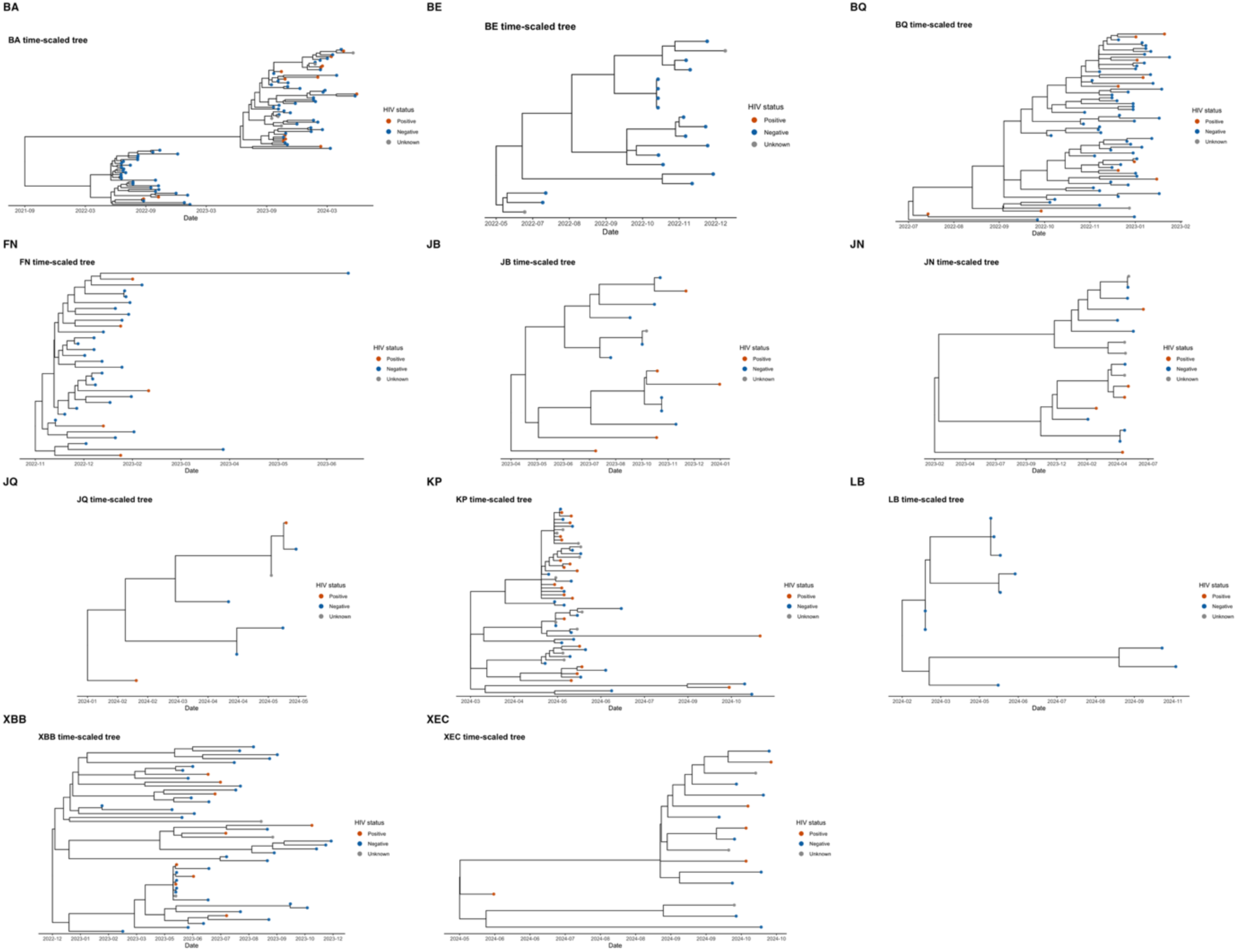
Lineage-specific time-scaled phylogenies of major SARS-CoV-2 lineages from Botswana (June 2022–November 2024), annotated by HIV status. Branch lengths are scaled to calendar time. Tip colors indicate HIV-positive, HIV-negative, or unknown status. These lineage-resolved trees depict within-lineage diversification and provide a framework to evaluate potential associations between HIV status and phylogenetic clustering.

## Discussion

We used phylogenetic analysis to describe the genomic epidemiology of SARS-CoV-2 during 2022 - 2024. We found three major groups shaping Botswana’s epidemic trajectory – the BA.4/BA.5 clade, the XBB lineages, and the BA.2.86/JN.1-derived clades – highlighting both their global evolutionary context and their local establishment through successive introductions. These findings are consistent with genomic surveillance across Africa during the same period. In South Africa, BA.4 and BA.5 rapidly replaced BA.2 in early 2022^16^, while studies from North^17^ and West Africa^18^ similarly documented frequent shifts in Omicron sublineages, reflecting both local diversifications driven by sustained community transmission and repeated importations associated with cross-border mobility. The subsequent dominance of XBB lineages in coastal Kenya (Kilifi) during November 2023-March 2024, together with ≥38 independent introductions followed by onward local transmission^19^, parallels our observations in Botswana, emphasizing the interconnectedness of regional transmission networks across Africa. Finally, the emergence and subsequent expansion of BA.2.86/JN.1-derived clades in Botswana from late 2023 through mid-2024 parallels global observations of highly immune-evasive Omicron subvariants that rose to dominance in early 2024^20^, with ongoing global spread thereafter^21^, highlighting the value of ongoing genomic surveillance to monitor regional and international transmission dynamics.

The three major lineage groups identified in Botswana represent key evolutionary phases in the later stage of the Omicron period. The BA.4/BA.5 clade, which emerged from the BA.2 backbone in early 2022, carried signature spike mutations such as L452R and F486V that enhanced viral transmissibility and conferred partial resistance to neutralizing antibodies^16^. These variants rapidly replaced BA.2 in southern Africa and subsequently spread worldwide^22^. The XBB lineages, first detected in late 2022, originated through recombination between two BA.2 sublineages (BJ.1 × BM.1.1.1), giving rise to highly immune-evasive recombinants such as XBB.1.5 and XBB.1.16^23^. These variants demonstrated stronger ACE2 binding affinity and broad antibody escape, driving large transmission waves globally and across Africa during 2023^19^. Subsequently, the BA.2.86/JN.1-derived clades represent the most antigenically divergent group, harboring more than 30 additional spike mutations relative to BA.2. These highly immune-evasive subvariants—exemplified by JN.1 and its descendants KP and XEC—rose to dominance from late 2023 through 2024^20^. Collectively, these lineages illustrate the ongoing adaptive evolution of SARS-CoV-2 through both recombination and convergent mutation, balancing enhanced transmissibility with progressive immune evasion.

Our findings suggest that HIV status was not a primary determinant of lineage diversification and did not shape the overarching phylogenetic structure. Nonetheless, subtle, localized effects on transmission cannot be excluded, and formal trait-association analyses are warranted to assess whether the limited same-status groupings observed exceed expectations under random mixing. Previous studies investigating the impact of HIV on SARS-CoV-2 evolution have largely focused on within-host dynamics and prolonged infection, particularly among individuals with advanced immunosuppression. For example, a case of persistent SARS-CoV-2 infection in a person with advanced HIV, where delayed viral clearance and prolonged replication led to the accumulation of adaptive spike mutations (e.g., E484K, K417T, N501Y) conferring antibody escape and promoting immune-evasive viral lineages^24^. More recently, Velasquez-Reyes et al.^25^ documented another prolonged infection lasting more than 750 days in an individual with uncontrolled HIV, showing parallel evolutionary trajectories in the spike and NSP6 regions resembling those in Omicron. However, population-level evidence linking HIV infection to broader patterns of viral diversification or transmission remains limited. Beyond HIV, age as another host characteristic has been analyzed for its effect on transmission dynamics. In a population-based modeling study, Davies et al.^26^ demonstrated that age-dependent differences in susceptibility and clinical presentation substantially shape SARS-CoV-2 transmission patterns, as children and adolescents are roughly half as susceptible to infection as adults and more likely to experience subclinical or mild illness, thereby contributing less to onward transmission in population-level models. Moreover, at the immunologic level, older individuals exhibit delayed immune activation and attenuated neutralizing antibody and T-cell responses following infection^27^, which may in turn influence viral clearance and onward transmission potential. However, few studies have formally incorporated host-level traits into phylogenetic analysis to quantify their contribution to transmission structure.

## Conclusion

This study of SARS-CoV-2 genomic epidemiology from the Greater Gaborone District revealed multiple introductions and six successive Omicron sublineage waves between mid-2022 and late 2024, indicating that Botswana’s epidemic was driven by repeated importations rather than sustained local diversification. Phylogenetic clustering of HIV-positive and HIV-negative sequences suggested that HIV infection did not markedly alter population-level diversification patterns, though localized dyads among HIV-positive hosts indicate potential within-lineage effects warranting further investigation. From a public health perspective, these results highlight the critical need to sustain genomic surveillance capacity in HIV-endemic regions, where immune heterogeneity and prolonged infections may foster viral diversification and immune escape. Future work should extend sampling to rural and underrepresented areas, conduct longitudinal follow-up of immunosuppressed individuals, and employ phylodynamic and transmission network approaches to better quantify how host-level immune variation shapes viral evolution and spread.

## Data Availability

All data produced in this study will be deposited in a public repository upon submission to a peer-reviewed journal.

## Data Availability

All data produced in this study will be deposited to a public repository upon submission to journal.

## Ethical Approval statement

The study received approval from the institutional review board at the University of California, Irvine, and the Botswana Ministry of Health and Wellness Human Research Development Committee. Written consent was obtained from all participants.

